# Health workers’ Perspective on the Feasibility and Acceptability of the Introduction of AgRDT for COVID-19 in Kisumu County, Western Kenya

**DOI:** 10.1101/2022.05.24.22275498

**Authors:** M. Omollo, I. A. Odero, H.C. Barsosio, S. Kariuki, F. Ter Kuile, S.O. Okello, K. Oyoo, A. K’Oloo, K. Otieno, S. van Duijn, N. Houben, E. Milimo, R. Aroka, A. Odhiambo, S. N. Onsongo, T.F. Rinke de Wit

**Author notes:** Corresponding Author, PO Box 1578-40100, Kisumu, Kenya.

## Abstract

COVID-19 pandemic remains a major global public health challenge also in Low- and Middle-Income Countries (LMIC), due to fragile health systems, limited resources and personnel, low testing and counseling capacity, community perceptions, among others. In Kisumu County of Western Kenya, a unique Public Private Partnership (PPP) was rolled-out to increase testing and capacity building by linking private facilities to the ongoing public sector efforts in combating COVID-19. It became increasingly clear that centralized PCR testing for COVID-19 was too labor-intensive, expensive, prone to machine breakdowns and stock-outs of essential reagents, resulting in long turn-around times and sometimes even adaptations of patient selection criteria. A clear need was identified for rapid point-of-care COVID-19 testing (AgRDT). After successful field evaluation, RDT for COVID-19 was offered through the PPP. This paper aimed to understand the health workers perspective on the feasibility and acceptability of the introduction of the AgRDT in Kisumu County.

In-Depth Interviews were conducted with selected health workers (n=23) from the participating facilities and analyzed using Nvivo 11 The health workers accepted the use of AgRDT as it enabled the strengthening of the existing health system, increased testing capacity and provided capacity building opportunities. Challenges included poor management of results discrepant with PCR gold standard.

The health workers applauded the introduction of AgRDT with the Kisumu County Department of Health as a more realistic and user-friendly approach, leading to fast turn-around times and increased personal safety experience.

## Introduction

The effect of COVID-19 in Low- and Middle-Income (LMICs) countries is manyfold, including socio-economic, political and health system-specific, such as shortages of health workers, inadequate commodities and consumables, difficult working circumstances to maintain Infection Prevention and Control (IPC) Measures (1). To increase health system capacity, a Public Private Partnership (PPP) was formed, named COVID Diagnostic Project (COVID-Dx) to assist the Kisumu County Department of Health (DoH) in combating COVID-19. This PPP is described elsewhere (2), but generally included increased testing capacity, digital data collection, timely reporting to patients and policy makers, and further capacity building of private facilities. The partnership was formed between PharmAccess Foundation (PAF), Kisumu County DoH and the Kenya Medical Research Institute/Centre for Global Health Research (KEMRI/CGHR).

During the implementation process of COVID Diagnostic Project, there was only one referencing laboratory in the region that was running the COVID-19 PCR (Polymerase Chain Reaction) tests. The PCR was considered the gold standard for COVID-19 diagnosis because of its accuracy and reliability (3). However, there are several challenges that came with the COVID-19 PCR testing. These included the need for expensive laboratory equipment, well-trained laboratory staff, costs of reagents and supply chain shortages (4). Other challenges include sample storage and sample transportation in (infectious) transport media (5). Moreover, PCR Tests Kits are expensive, have relatively long turn-around time, need highly trained laboratory technicians (6). The use of rapid tests, such as COVID Antigen Rapid Diagnostic Testing (AgRDT) became handy in LMICs as it minimized turnaround time, did not require (cooled) sample transportation to the reference laboratory, and needs minimal laboratory technical training (7).

Due to challenges experienced during the rollout of the COVID-Dx PCR testing, a decision was made to conduct a field evaluation of the AgRDT that showed promising evaluations in other setups (7). In Kenya no empirical information was available on AgRDTs and the perceptions around usage. This paper reports on Kenyan health care workers’ perspectives on feasibility and acceptability of COVID-19 AgRDT. The study was conducted in selected private and public health facilities within Kisumu County of western Kenya under the PPP COVID Diagnostic Project.

## Methods

### Study Setup and Population

The PPP is a collaboration between Kisumu County DoH, Kenya Medical Research Institute/Center for Global Health Research (KEMRI/CGHR) in Kisumu, PharmAccess Foundation and selected healthcare facilities in Kisumu East and Central Sub counties. Health care workers were interviewed to get their perspectives on the use of AgRDT. There were six participating health facilities both private and public (Table 1)

**Table 1:** Study Participants for IDI

| **Health Facilities** | **Health Workers** | **Classification** |
| --- | --- | --- |
| A | 4 | Private Health Facility |
| B | 4 | Public Health Facility |
| C | 3 | Public Health Facility |
| D | 4 | Private Health Facility |
| E | 4 | Private (Mission) Health Facility |
| F | 4 | Private (Mission) Health Facility |
| Total | 23 |  |

### Study design and procedures

Research design adopted the Grounded Theory qualitative design. In-Depth (IDIs) Interviews were used for data collection. A total of 23 health workers were selected from the 6 participating facilities consisting of 2 public and 4 private health facilities: 2 of them being run under the Mission. The criteria for choosing the participating facilities were mainly categorized into two: essential and non-essential criteria. The essential criteria for private facilities include: provider should have a license, permission by DoH to participate in the COVID-AgRDT project, reasonable patient throughput and proven ability to detect positive COVID-19 cases, a working and serviced fridge and generator for sample storage, willingness of qualified lab staff to be trained for AgRDTs and willingness of management to participate in COVID-AgRDT project. The criteria for public facilities was based on those proposed by DoH to participate in the study. Some considerations for participation of proposed public hospitals included referral and county hospitals with reasonable patient throughput and proven ability to detect positive COVID-19 cases, willingness of qualified lab staff to be trained for AgRDTs and willingness of health facility in-charges to participate in COVID-AgRDT project Purposive sampling method was applied for the selection of participants for the IDIs by both study staff and facility-based team leads. The health workers consisted of laboratory technicians, clinical officers, nursing officers, and psychosocial counsellors who are involved in COVID-19 related activities in the health facilities.

### Protection of human participants

Selection of participants were done using purposive sampling technique. Interview appointments were made by the study staff through both emails and phone calls. Consent to contact the participants was obtained verbally. Thereafter, written consents and participants’ demographic information were obtained prior to conducting of the interviews.

The interviews were conducted within the health facilities on varying locations like private rooms with observance of confidentiality and privacy codes. The benefits of participation in the study was contributing to the evaluation of the AgRDT kit. In addition, there were provision of transport reimbursement to ease the burden of expenditure from the participant. The risk were participants not being comfortable o respond to some of the question which was mitigated by giving them options to respond to the questions they were comfortable with.

### Ethical approvals and consent to participate

The Jaramogi Oginga Odinga Teaching and Referral Hospital (JOOTRH): Institution and Ethical Review Committee provided research and ethical approval with license number IERC.IBlVOL.tt/3SS/20. Additional research and ethical approval was provided by the National Commission for Science, Technology and Innovation (NACOSTI) with License Number ABS/P/20/7959. All participants and selected health facilities provided written consents to participate in the study

### Data collection and analysis

Data collection was done by trained interviewers using open-ended interview guide which was designed to elicit responses on Private Public collaborative responses, and experiences with COVID AgRDT services as a way of getting their perspective on its feasibility and acceptability. The interviews were conducted in either English, Swahili or Luo. Data from the IDIs were audio recorded and transcribed in a verbatim form. Proof reading was done on the transcribed data to ensure quality transcription. Interpretation of the data was done thematically using Nvivo 11 software. This involved the development of the codebook and the scripts imported into Nvivo 11. Coding was done by two independent coders with intercoder reliability performed followed by generation of reports. Quality checks right from data collection to data analysis were appropriate in terms of credibility and confirmability.

## Results

Demographic characteristics of the respondents was captured in terms of gender, education level, years of experience in the current position (Table 2). Of the respondents interviewed for the AgRDT feasibility and acceptability study, half were male, with 87% having completed at least diploma education. In terms of professional cadre, 4 were nursing officers, 6 were clinical officers, 9 Lab technologists and 6 were psychological counselors (Table 2).

**Table 2.** Demographic characteristics of the respondents.

|  |  |  |
| --- | --- | --- |
| Gender |  |  |
|  | Male | 12 |
|  | Female | 11 |
| Age |  |  |
|  | 20-30 | 8 |
|  | 31-40 | 11 |
|  | 41 and above | 4 |
| Education |  |  |
|  | Certificate | 3 |
|  | Diploma | 11 |
|  | Bachelor's Degree and above | 9 |
| Cadre |  |  |
|  | Nursing Officers | 4 |
|  | Clinical Officers | 4 |
|  | Laboratory Technologists | 9 |
|  | Psychosocial Counsellors | 6 |

Healthcare workers’ (HCW) perspectives on the feasibility and acceptability of AgRDT were explored with respect to staff workload, availability of test kits, contact tracing, acceptability of AgRDT, referrals, decentralization versus centralized lab PCR, cost implications to HCWs and community, challenges with using AgRDT (discrepant results) and testing procedures.

### Staff workload

On the aspect of staff workload, the healthcare workers, both public and private were contented with AgRDT’s capacity to reduce patient congestion and staff work overload. Through the adoption of AgRDT these facilities were now able to provide customer-friendly COVID AgRDT services.

“*We complement each other. So, it is about having as system where when we are overwhelmed, we can send patients to them (To mean public facilities) and they can send patients to us. This happens especially in this pandemic like where our ICU capacity is quite limited. So, where we don’t have that capacity, we transfer some patients to them and vice versa*.*” IDI 006 A, H/W 4*

### Availability of kits

In addition, both sectors complemented each other through borrowing test kits in cases where either of the participating health facilities had experience a high turnover of test demands.

The usage of AgRDT made it possible to have activities around Contact tracing, referrals in cases where the isolation and quarantine rooms were fully occupied easily operational.

*“We also have same catchment areas, so we more or less experience same challenges in COVID. Sometimes we get overwhelming numbers, so we refer our patients or transfer to them (To mean public health facilities)*.*” IDI 006 A, H/W 4*

### *Healthcare system*s

The introduction of the usage of the AgRDT kits came with a well-received capacity building of both private and public health facilities, involving provision of PPEs, on-the-job sample collection and testing training and interpretation of AgRDT kit results.

*“Yeah, we exchange knowledge; because you can consult where you don’t understand. And we also exchange materials; in-case we don’t have enough materials; we can borrow from the public. And then in cases that we maybe have a special case that needs to be done like urgently, we sometimes refer to the public*.*” IDI 016 F, H/W 12*

The pre- and post-test counselling that was offered to the patient during the COVID AgRDT service provision helped with preparing and informing the patient of what was expected throughout the process. The role of the psychosocial counsellor was considered key, both for motivating a patient to get tested and later during the delivery of the patient results.

*“My take is that we need, like what was lacking, we need to make sure that the patient is well informed because they come in for now testing. Like the time of counselling, they need to be told. You know like what was coming out the time of taking the sample, now you have to explain again to the patient. But I was thinking if the patient is well informed on what is going to be done during sample collection, I think they could understand more and cooperate well*.*” IDI 26 E, H/W 20*

### Cost implications

The introduction of the usage of COVID AgRDT kit came with relief on the cost of conducting COVID-19 test. The private facilities participating in the study were not allowed to ask for any charges of the COVID-19 testing, only the general consultation fees an experience which went well. The respondents noted that the method was cheap, and affordable as compared to the COVID19 PCR testing. In addition, it required no machines and electricity and it could be performed in-house at the point-of-care.

*“It is cheaper as compared with the PCR. You know PCR needs the machine with you, the person, though the RDT you also need the person. For PCR you need the machine itself, the machine will need a reagent to run it and then let me say when the machine breaks down you have to maintain the machine. But the RDT you don’t need to maintain the machine*.*” IDI 02 B, H/W 2*

*“I think it looks fairly easy to use because with minimal trainings, you don’t need any other additional machines to use it because it has an in-built control. And so, there’s no need for transportation of samples. Certainly, it’s much better because we can do it in-house, and we can do it bedside*.*” IDI 06 A, H/W 04*

### Challenges in discrepant results

However, the health workers noted scenarios with discrepant results when confirmatory COVID-19 PCR test was done. Although the operating procedures of AgRDT clearly included informing the participants about the pro’s and the cons of a rapid test, this not always transpired. In particular (false-)negative AgRDT results, followed the next day by a positive PCR created confusion.

*“It is not easy. Because at first, let’s say the RDT is positive, then you have counselled this patient and you have told him this one is positive and then the following day you give him another different report. So it is not something normal. But that is not bad as such. The difficult part comes when the RDT is negative, and then the PCR comes as positive. Convincing somebody that this result is a bit different, he starts querying: Are we doing the right thing, or what is wrong?” IDI 002 B, H/W* 2

### Communication challenges

The respondents noted lack of proper communication channels to reemphasize the standard operating procedures of sample collection. Some facilities were collecting single nasal pharyngeal swabs while others were collecting double nasal pharyngeal swabs; some facilities first did PCR sampling and then AgRDT, others the other way round. This brought some differences in results and concomitant misunderstandings.

*“As far as we have gone with this COVID-19 program, the only challenge that we are experiencing is miscommunication and that was only in the first stages. For example, some facilities were not doing the procedure the right way. We saw that as a challenge. Like some facilities were not getting enough in terms of capacity building*.*” IDI 07 A, H/W 5*

*“Another challenge is communication barrier. Sometimes you can make a phone call, you find somebody at the public facility maybe is not in, will refer you to another person. So, you’ll go round before you get assistance as maybe you required*.*” IDI 21 D, H/W 16* Some respondents noted that the consenting process was good, however it was long and very much time consuming.

*“Actually, you had to take the client through the session and then the client consent and then that is when the client is ready to be taken for the sample. So I can also say this is another challenge, because it takes a lot of time. Just taking the client through all these until the client accepts and understand that is when you will be able to go on” IDI 17 F, H/W 13*

## Outcome of the usage of AgRDT

Health workers noted increase in their knowledge of COVID AgRDT services, as a result of the various trainings that the project provided in addition to the increased interaction between private and public facilities including mentorship during supportive supervision.

*“They share knowledge, interactions and the experiences since the public tend to have more than them. Because in private people know that you must go there with cash making the public sector to receive more patients. So when they interact with the patient’s they tend to gain more experience. So those people might come and borrow these experiences and challenges which we are undergoing so I think through that one they can get a lot from that*.*” IDI 02 B, H/W 02* Increased number of patients were able to access COVID AgRDT services at either the private or the public facilities and this enabled faster ways of screening for COVID-19.

## Discussion

In the early phases of the epidemic, there was simply no access to RDTs for the Kenyan private sector. RDTs were only entering the country through international development aid. However, more than half of healthcare in Kenya is delivered through the private sector and therefore we considered they should also be facilitated to have access to RDTs. The RDT tested in our PPP project was of internationally recognized quality, could be ordered easily at lower quantities-suitable for individual hospitals, at low price (<$3/test) and shipped from South Korea to Nairobi within a week. Therefore, this test was chosen to be field-evaluated, which actually was accomplished (8). Currently the process has been started to license this AgRDT for use in Kenya. The current feasibility and acceptability study was performed amongst (private) healthcare workers to assess the potential of such a test for broader usage.

The study gave insight of the health workers’ perception on the feasibility and acceptability of the introduction of AgRDT for COVID-19 testing. Feasibility appeared realistic, given the simple testing procedures. Testing cues were successfully combated with AgRDT preventing congestions. Expansion of testing sites, availability of resources and provision of technical expertise were some of the perspective of the health workers that enabled the feasibility and acceptability of the introduction of AgRDT (9). The increase in testing capacity came handy as that was the same period the Delta variant was spreading fast in Kisumu County. Collaborative responses through PPPs have always worked hand in hand to achieve expanded capacity in service delivery both clinical and non-clinical, through the provision of commodities and equipment (10).

Furthermore, this study showed that an overwhelmed COVID-19 testing system fully relying on the usage of PCR was facing numerous challenges such as delays in turnaround time that was affecting patient management and subsequent spread of the epidemic (4). Therefore, the introduction of the AgRDT was considered a much better option to the PCR, as it offered point of care service and efficiency in terms of faster delivery of results (11). Other benefits of the AgRDT included fairness in terms of cost of operations as it did not require technical manpower, cost of transportation to the reference lab and cost of running the sample (12).

In addition, the numerous trainings, mentorship and supportive supervisions that were offered to the health workers during the implementation of the study, increased their knowledge on the provision of COVID-19 testing services. Moreover, there were increased number of patients who could now have access to COVID-19 testing as a result of the introduction of the AgRDT which facilitated a significant scale up in COVID-19 testing (13)

Furthermore, the role of psychosocial counsellor was key during the project. The counsellor offered Pre and Post-Test counselling to the patients. This is because COVID-19 can have a great negative impact on mental health and psychosocial life (14).

There were two limitations that the health workers noted about the study. First, the study was designed as a pilot to the Kisumu County Department of Health, in which the usage of AgRDT was done alongside confirmatory PCR testing. Although participants were informed about the possibility of discrepant results, these did result in some confusion. It was found out that a major cause of the discrepancy was attributed to non-adherence by healthcare staff to the study procedures where sometimes one and the same nasal pharyngeal swab was used for both AgRDT and PCR testing. This could result in false-negativity, due to dilution of the sample (15). Secondly, the test-result communication channels were not always clear and changed during the project, involving 3 different partners in addition to the participating facilities. There were no formal DoH protocols on communication channels, and in some cases feedback was not relayed to patients in a timely manner causing frustration and anxiety (16).

## Conclusions

The introduction of the rapid AgRDT in Kisumu was considered a success by healthcare workers in improving the COVID-19 service provision in the selected health facilities. The Kisumu County DoH was able to escalate the usage of AgRDT to non-participating health facilities. Future policy should include up-to-date and clear communication channels of COVID-19 test results.

**Figure 1:**
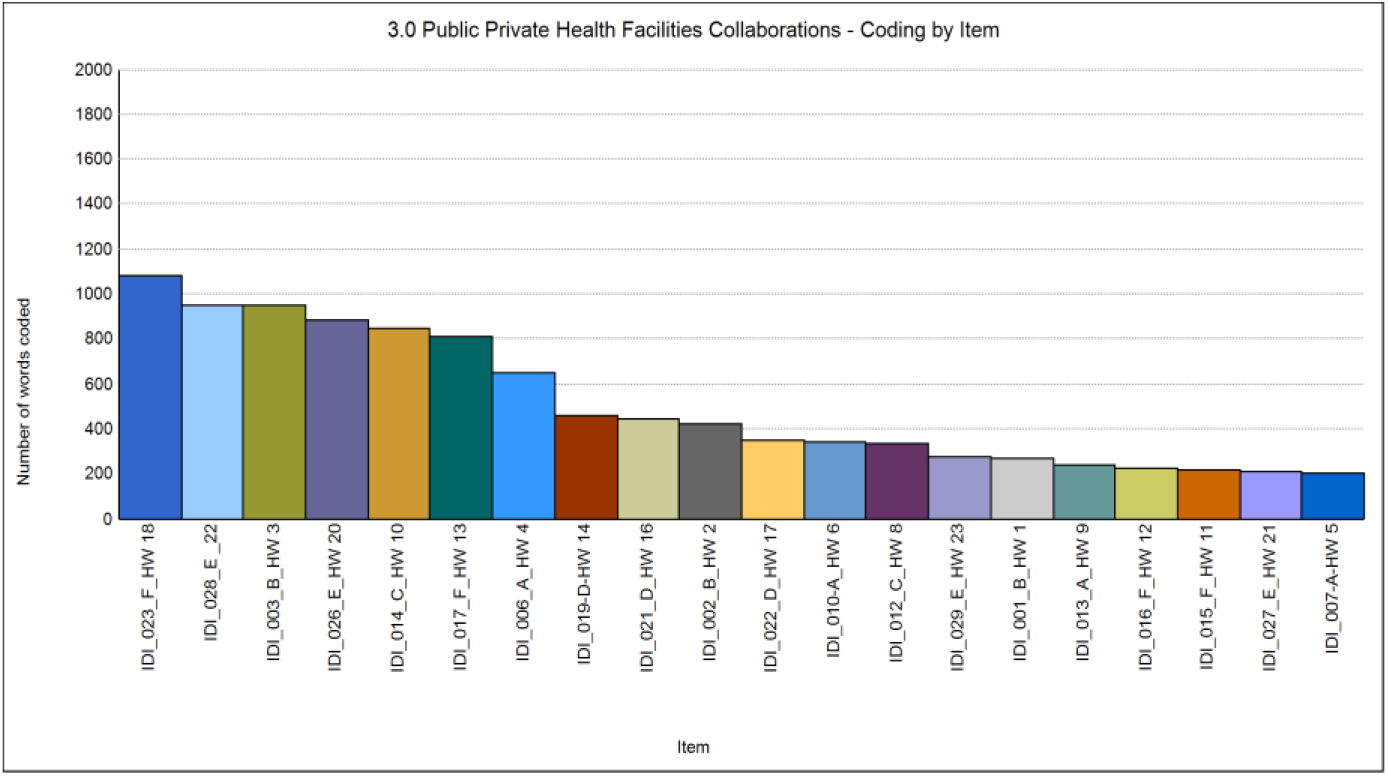
Coding by item. Number of words coded on Public Private Health Facilities Collaborations. IDI 023 shared the most on the theme during the interviews.

## Data Availability

All relevant data are within the manuscript and its Supporting Information files

## Acknowledgments

We give thanks to the PharmAccess Foundation for technical advice throughout the implementation process, to KEMRI/CGHR for running confirmatory PCR test and to the Kisumu County DoH for taking its lead role in facilitating the study.

## Notes

### Competing Interest Statement

The authors have declared no competing interest.

### Funding Statement

The statement should include: • Specific grant numbers. This research was supported by a grant from Achmea Foundation April 15, 2020, grant number 2020.002. PharmAccess is supported by the Netherlands Ministry of Foreign Affairs. • Initials of authors who received each award T.F. Rinke de Wit • Full names of commercial companies that funded the study or authors Not Applicable • Initials of authors who received salary or other funding from commercial companies Not Applicable • URLs to sponsors’ websites https://www.achmea.nl/en/foundation/ https://www.government.nl/ministries/ministry-of-foreign-affairs Also state whether any sponsors or funders (other than the named authors) played any role in: • Study design KEMRI/CGHR and PharmAccess Foundation developed the research protocol of the study • Data collection and analysis Not Applicable • Decision to publish Not Applicable • Preparation of the manuscript Not Applicable

### Author Declarations

Ethical approvals and consent to participate The Jaramogi Oginga Odinga Teaching and Referral Hospital (JOOTRH): Institution and Ethical Review Committee provided research and ethical approval with license number IERC.IBlVOL.tt/3SS/20. Additional research and ethical approval was provided by the National Commission for Science, Technology and Innovation (NACOSTI) with License Number ABS/P/20/7959. All participants and selected health facilities provided written consents to participate in the study

